# APOLLAI, a multimodal AI system, versus EMS personnel for prehospital detection of stroke-related deficits

**DOI:** 10.64898/2026.10.16.26365784

**Authors:** Cédric Javault, Estefania Vargas Gonzalez, Lionel Pibre, Félix Yriarte, Eric Thouvenot

**Author notes:** Corresponding author E-mail addresses. These authors contributed equally (Cédric Javault, Estefania Vargas Gonzalez).

## Abstract

**Background and Purpose:** Prehospital stroke recognition remains suboptimal, with nearly one-third of strokes unidentified before hospital arrival, and neurological assessment being inherently subjective, with substantial interobserver variability even among certified clinicians. APOLLAI, a multimodal artificial intelligence system, was developed to detect stroke-related neurological deficits from video and audio recordings. We compared the diagnostic performance of APOLLAI and emergency medical services (EMS) personnel against an expert consensus reference standard.

**Methods:** In this prospective study, video recordings of 295 patients with suspected stroke (mean age, 70.3±13.3 years) were obtained at Nîmes University Hospital (August 2023–July 2025). Five National Institutes of Health Stroke Scale items (best gaze, facial palsy, motor arm, aphasia, dysarthria) were independently annotated by seven certified stroke neurologists, combined into consensus labels using the Dawid–Skene algorithm, and separately assessed by EMS personnel (mean of 15 raters per patient). APOLLAI, a multimodal machine-learning framework that combines computer vision, audio-based deep learning, and a final support vector machine (SVM) fusion classifier, was evaluated against EMS using sensitivity, specificity, and the area under the receiver operating characteristic curve (AUC).

**Results:** Interrater agreement among the seven neurologists was moderate overall (mean κ=0.541; range, 0.367–0.732), indicating an intrinsic difficulty of neurological assessment. APOLLAI showed higher sensitivity than EMS for all five neurological tasks and for the global assessment (78.5% versus 59.4%), with broadly comparable specificity (87.0% versus 85.9% for the global assessment). AUC values ranged from 0.839 (facial palsy) to 0.966 (best gaze), with a global AUC of 0.882.

**Conclusions:** APOLLAI demonstrated higher sensitivity than EMS personnel for detecting stroke-related neurological deficits, with preserved specificity. These findings support its potential as a standardized, scalable adjunct to prehospital stroke recognition, warranting prospective validation under real-world EMS conditions.

## Introduction

Stroke is a leading cause of death and long-term disability worldwide, affecting approximately one in four adults during their lifetime and causing death or permanent disability in nearly 60% of cases^1–3^. Its global burden continues to rise despite advances in prevention and acute management^4^ and also translates into substantial economic costs, estimated at 3% to 4% of total healthcare expenditure in Western countries, with projected costs in the United States alone rising from $105.2 billion in 2012 to $240.7 billion by 2030^5^. Early treatment remains the strongest modifiable determinant of outcome, as every minute of untreated cerebral ischemia is estimated to destroy 1.9 million neurons^6^. The biological urgency is embodied in the maxim “Time is Brain”, and reinforces the imperative for rapid stroke recognition and expedited access to acute stroke care.

In this context, emergency medical services (EMS) play a central role in the early identification, triage, and transport of patients with suspected stroke. Rapid recognition of stroke symptoms, hospital prenotification, and appropriate triage to specialized stroke centers are essential components of modern stroke systems of care, and treatment guidelines emphasize coordinated systems aimed at minimizing the time from symptom onset to treatment. Despite this pivotal role in the stroke chain of survival, prehospital stroke recognition remains suboptimal, with nearly one-third of patients with stroke not identified before hospital arrival^7–9^, potentially delaying access to reperfusion therapies and reducing opportunities for favorable neurological outcomes. Prehospital stroke recognition depends primarily on the clinical assessment of neurological deficits, a process inherently prone to interobserver variability. Stroke symptoms are often heterogeneous, subtle, or fluctuating, and their identification depends on the examiner’s ability to recognize abnormalities. As a result, diagnostic accuracy may vary substantially across individuals, contributing to persistent under-recognition of stroke in the prehospital setting.

In prehospital emergency care, EMS personnel routinely rely on objective, device-based measurements to assess critical conditions including cardiac monitoring, pulse oximetry, blood glucose testing, and blood pressure measurement providing standardized, reproducible data independent of examiner expertise. No equivalent objective measurement tool exists for the assessment of stroke-related neurological deficits. To address this gap, numerous simplified clinical examination-based stroke scales have been developed and are widely used by EMS personnel, including the Face Arm Speech Time (FAST) scale^10^, the Cincinnati Prehospital Stroke Scale (CPSS)^11^, the Los Angeles Motor Scale (LAMS)^12^ and others. The National Institutes of Health Stroke Scale (NIHSS)^13^, although more comprehensive, is primarily administered by trained neurologists and stroke specialists in hospitals rather than EMS personnel in the field. Unlike cardiac or metabolic parameters, neurological assessment relies entirely on clinical observation and human interpretation, rendering it inherently subjective, highly dependent on examiner training, and subject to substantial interobserver variability even among certified clinicians.

Various technology-assisted approaches including near-infrared spectroscopy, ultrasound-based technologies, electroencephalography, microwave devices, and volumetric impedance spectroscopy have also been investigated^14^. However, most of these approaches require specialized equipment and trained personnel, which limits their scalability and routine implementation. Median acquisition times of approximately three to five minutes have been reported for several of these technologies^14^.

To address the persistent challenges of prehospital stroke recognition and the variability inherent to neurological assessment, we developed APOLLAI, a multimodal artificial intelligence system designed to detect stroke-related neurological deficits from brief video and audio recordings of the patient. In this study, we compared the diagnostic performance of APOLLAI and EMS personnel for the detection of key neurological abnormalities relevant to stroke recognition, using expert consensus annotations derived from seven certified stroke neurologists as the reference standard. We hypothesized that APOLLAI would demonstrate higher sensitivity than EMS personnel for the detection of individual neurological deficits while maintaining comparable specificity. We further assessed interobserver variability among expert neurologists and evaluated APOLLAI performance within the context of human expert disagreement, to explore its potential as a standardized and scalable tool for prehospital stroke assessment and triage.

## Methods

### Study Design

This study was conducted in four phases: 1. A prospective data collection phase, during which video recordings of patients admitted at the stroke unit for suspicion of stroke were obtained at Nîmes University Hospital (CHU Nîmes), 2. An annotation phase, during which these recordings were independently evaluated by expert and NIHSS-certified neurologists, a consensus vote being computed using Dawid-Skene algorithm^15^, 3. A training phase of the AI models and 4. An evaluation phase in which both EMS personnel and APOLLAI assessed the annotated video and performances against the ground truth were compared. The fourth phase of the study was developed in collaboration with the Service Départemental d’Incendie et de Secours de l’Hérault (SDIS34), the regional emergency medical response service responsible for approximately 1.26 million inhabitants in southern France; over 2,000 staff participated. The study protocol was approved by the ethics committees (CESREES file number 10625271 and 19715357 – CPP file number 2022-A01528-35) and the French National Commission on Informatics and Liberty (CNIL deliberations 2023-053 and 2023-054 + decision DR-2025-014).

### Outcome Measures

The primary outcome for phase 2 was the inter-rater agreement (measured by Cohen’s Kappa coefficient^16^) among the seven expert annotators in assessing the NIHSS score using videos of the patient performing the tasks corresponding to item 2 (best gaze), 4 (facial palsy), 5 (motor arm), 9 (best language / aphasia) and 10 (dysarthria). Secondary outcome was the evaluation of each neurologist against the ground truth from the six other neurologists.

The primary outcome for phase 4 was the diagnostic performance of APOLLAI and EMS personnel (measured by sensitivity and specificity) for the detection of neurological abnormalities corresponding to the five selected NIHSS items and for global stroke assessment, against the ground truth computed using the Dawid-Skene algorithm^15^ to compute a consensus vote among the seven neurologists. Secondary outcomes were the receiver operating characteristic (ROC) analysis and area under the curve (AUC)^17,18^ for APOLLAI.

### Phase 1: prospective data collection phase

A structured video dataset was prospectively created at Nîmes University Hospital (France) between August 2023 and July 2025 from participants with suspected stroke and healthy controls, recorded during the complete assessment of the NIHSS protocol. Among others, videos of each patient were acquired with the participant performing the following NIHSS tasks: best gaze (item 2), facial palsy (item 4), motor arm (item 5), best language (item 9), and dysarthria (item 10).

A total of 300 patients with suspected stroke were enrolled; two were subsequently excluded because of unusable video recordings and three withdrew consent, leaving 295 patients with complete data available for analysis. The patients ranged in age from 24 to 98 years (mean age, 70.3 ± 13.3 years), and 36.9% were female. Demographic and clinical characteristics, including age, sex, and final diagnosis, were systematically collected for all participants. Final diagnoses were distributed as follows: 248 ischemic strokes, 16 hemorrhagic strokes, 26 transient ischemic attacks, and 5 stroke mimics.

The final dataset comprises 295 patients with suspected stroke and 85 demographically matched healthy controls. In addition, 55 patients were filmed again several months after the acute event, once most or all neurological deficits had resolved.

### Phase 2: Annotation phase and Consensus vote scheme

A panel of seven expert stroke neurologists, all certified in the National Institutes of Health Stroke Scale and routinely performing NIHSS-based assessments in daily clinical practice, independently annotated the entire dataset of 435 subjects on the following five NIHSS items from video recordings: best gaze (item 2), facial palsy (item 4), motor arm (item 5), best language (item 9), and dysarthria (item 10). These symptom-level annotations were used as ground truth for model training and evaluation, defining the reference standard as the examining neurologist’s bedside clinical judgment on each NIHSS item rather than a computed tomography (CT) or magnetic resonance imaging (MRI)-based diagnosis. APOLLAI was accordingly designed to approximate this clinical assessment, not to replace neuroimaging.

To generate a robust reference standard from expert annotations, consensus labels were computed using the Dawid–Skene probabilistic algorithm^15^. This method estimates latent true labels while accounting for annotator-specific variability and biases.

One-versus-rest (OVR) analyses were also performed, where consensus labels were recalculated using six neurologists while excluding the neurologist being evaluated. The excluded neurologist’s performance was then assessed relative to the consensus established by the remaining six annotators.

Inter-rater agreement among expert annotators was assessed for each individual task and for the global assessment using Cohen’s κ coefficient^16^ for pairwise comparisons and summarized across all annotator pairs. Agreement was categorized as slight, fair, moderate, substantial, or almost perfect according to standard benchmarks^19^.

### Phase 3: APOLLAI Model Development

APOLLAI was developed as a multimodal machine learning framework designed to detect stroke-related neurological deficits from synchronized video and audio recordings. Separate task-specific models were trained to identify neurological abnormalities corresponding to each of the five selected NIHSS items, using the consensus vote of phase 2 as ground truth.

For the assessment of best gaze (item 2), facial palsy (item 4), and motor arm impairment (item 5), video recordings were preprocessed using computer vision algorithms which were subsequently fed into machine learning algorithms/processes. Audio tracks extracted from video recordings were preprocessed prior to classification using deep learning models for assessment of aphasia (item 9) and dysarthria (item 10). Predictions from individual task-specific models were subsequently integrated through a fusion classifier based on a SVM^20^ to generate a global APOLLAI result. Detailed technical specifications regarding model architecture, feature extraction, and training procedures have been described previously^21^.

To assess model robustness and generalizability, a repeated 5-fold cross-validation strategy was employed. The dataset was randomly reshuffled 25 times, and for each repetition, a 5-fold cross-validation was performed. At each fold, 80% of the patients were allocated to training and validation, while the remaining 20% were used for testing. Within the training and validation set, 20% of the patients were further reserved for validation, resulting in approximate training, validation, and test proportions of 64%, 16%, and 20% of the overall dataset, respectively.

All data partitioning was performed at the patient level to prevent information leakage. Consequently, all recordings from a given individual, including multiple videos and longitudinal acquisitions collected several months apart, were always assigned to the same subset within a given split. Overall, this procedure yielded 125 trained models (25 x 5 folds).Phase 4: Evaluation of EMS and APOLLAI against the ground truth

EMS diagnostic performance was evaluated prospectively by approximately 2,000 firefighters of *“Service Départemental d’Incendie et de Secours de l’Hérault” (SDIS34)*, during mandatory continuing professional education sessions held between February and June 2025. Many sessions were organized, each of them regrouping approximately 10 EMS personnel. During each session, the group was shown video recordings of six patients from the phase 1 dataset. Participants used their smartphone and a QR code displayed alongside the videos to access a dedicated web platform and independently evaluate each patient. For each patient, each EMS personnel was first asked whether the patient exhibited clinical signs of stroke. If a participant answered positively, they then had to specify the neurological tasks in which abnormalities were observed. Each of the 288 patients (some patients’ videos were not available at the time) was evaluated for the five tasks by a mean of 15 EMS personnel.

Because not all patients in the dataset had been evaluated by the same number of EMS personnel, for EMS assessment, predictions were normalized at the patient level to ensure equal contribution from each patient.

The ground truth was the NIHSS score for each task ranging from 0 to 2 for eye deviation and dysarthria, from 0 to 3 for facial palsy and best language, and from 0 to 4 for motor arm. EMS assessment was not an NIHSS score but a Normal / Abnormal answer for each task, and, for this study, APOLLAI was trained as a binary classifier outputting ‘Normal’ or ‘Abnormal’ for each task, the classification threshold being selected as the value maximizing the Youden index over the test set^22^. NIHSS scores were therefore binarized to assess EMS and APOLLAI’s performances (with NIHSS = 0 corresponding to ‘normal’, and NIHSS ≥ 1 corresponding to ‘abnormal’). Diagnostic performance was then evaluated using sensitivity, specificity for APOLLAI and EMS, and the AUC for APOLLAI.

Global stroke assessment for EMS and neurologists was defined as abnormal if and only if one task was assessed as abnormal (and global assessment was normal if all tasks were assessed as normal); global assessment for APOLLAI was defined as the output of a fusion classifier based on a SVM, which integrated predictions from all five individual neurological tasks into a single global score.

For each task and for the global assessment, EMS and APOLLAI performances were further benchmarked against individual neurologist performance using an OVR approach, in which each neurologist was successively excluded and their diagnostic performance assessed relative to the consensus established by the remaining six annotators. For each of the seven resulting ground-truth configurations, a full model was retrained (125 models per configuration, corresponding to 25 repetitions of 5-fold cross-validation), yielding closely similar ROC curves across configurations. For clarity, the ROC curves and the EMS performance points reported in the Results correspond to the model trained on the consensus of all seven neurologists; the seven ROC curves and EMS performance points corresponding to the OVR approach are presented in the supplementary materials.

All statistical analyses were performed using Python (version 3.12.12; Python Software Foundation, Wilmington, DE, USA) with scikit-learn (version 1.8.0).

## Results

### Inter-rater agreement along NIHSS tasks

Inter-rater agreement among the seven certified neurologist annotators was assessed for each of the five neurological tasks individually and for the global assessment as defined in the previous section. Task-level pairwise Cohen’s Kappa values (κ) are presented in Figure 1 and summarized in Table 1 using minimum, maximum, and mean value for each task.

**Table 1.** Interobserver agreement across the five neurological assessments. Cohen’s κ values (minimum, mean, maximum, and standard deviation) for each neurological task.

|  | Min | Average | Max | Standard deviation |
| --- | --- | --- | --- | --- |
| Best gaze | 0.17 | 0.437 | 0.665 | 0.165 |
| Facial palsy | 0.219 | 0.422 | 0.544 | 0.076 |
| Motor arm | 0.548 | 0.714 | 0.81 | 0.088 |
| Aphasia | 0.272 | 0.494 | 0.608 | 0.095 |
| Dysarthria | 0.372 | 0.547 | 0.665 | 0.084 |
| Global | 0.367 | 0.541 | 0.732 | 0.102 |

**Figure 1.**
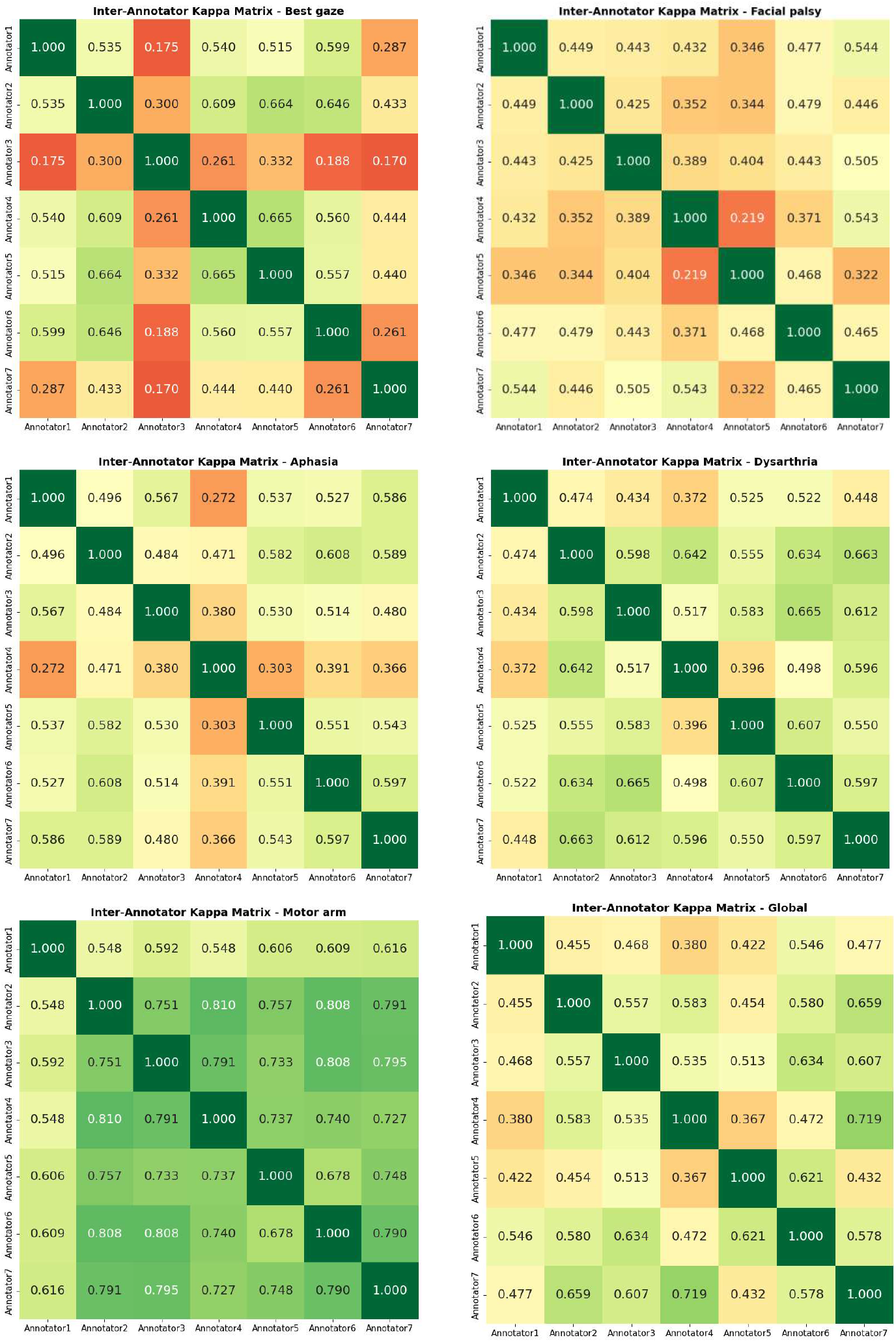
Interobserver agreement matrix among certified neurologist annotators. Heatmap showing pairwise Cohen’s κ coefficients among the seven certified neurologist annotators, for each of the five evaluated NIHSS items (best gaze, facial palsy, motor arm, aphasia, and dysarthria) and for global assessment.

Task-level agreement among the seven certified neurologist annotators demonstrated variability across neurological items. Agreement was highest for motor arm assessment (mean κ=0.714, substantial agreement; SD=0.088; range: 0.548 to 0.810), followed by dysarthria (mean κ=0.547; SD=0.084; range 0.372 to 0.665) and aphasia (mean κ=0.494; SD=0.095; range 0.272 to 0.608), both reflecting moderate agreements. Best gaze showed a similarly moderate mean agreement (κ=0.437) but the widest dispersion across annotator pairs (range 0.170 to 0.665), reflected by the highest standard deviation (SD=0.165) and the minimum value reaching the slight agreement range (κ=0.170). In contrast, facial palsy (mean κ=0.422; range 0.219 to 0.544) showed the most consistent agreement across annotator pairs, with the lowest standard deviation (SD=0.076) despite the lowest mean κ value of 0.422 corresponding to a moderate agreement. For the global assessment, the inter-rater agreement ranged from 0.367 (annotators 5 and 4) to 0.732 (annotators 5 and 7) with a mean value of 0.541, reflecting moderate agreement.

### Individual performance of the neurologists in the OVR approach

As all neurologists contributed to the main ground truth computed with a consensus vote of the seven neurologists, we used the OVR approach to assess individual performance. Results in the terms of sensitivity and specificity of the seven neurologists in the OVR approach are presented in Table 2.

**Table 2.**
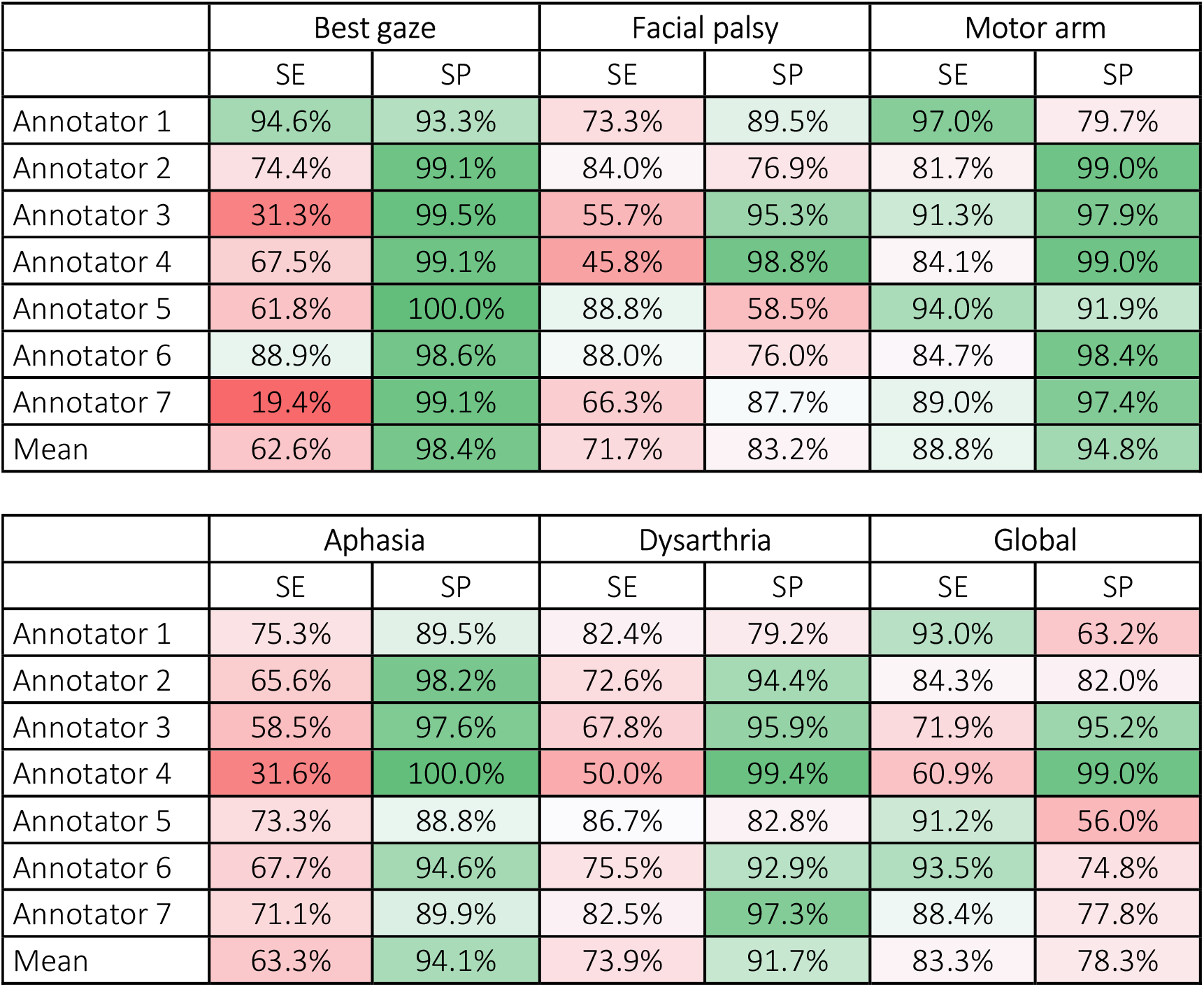
Individual performances of the seven annotators in the OVR approach. Sensitivity (SE) and Specificity (SP) are computed for each neurologist using the consensus vote of their six colleagues, at the task level and for global assessment.

Although specificity was high across tasks, with mean values ranging from 83.2% (facial palsy) to 98.4% (best gaze), sensitivity was lower and showed greater variability, particularly for best gaze (19.4%-94.6%), aphasia (31.6%-75.3%), and facial palsy (45.8%-88.8%). At the global assessment level, consistent with the moderate agreement observed, none of the certified neurologists demonstrated both sensitivity and specificity above 80%. For example, annotator 4 had 99.0% in specificity but only 60.9% in sensitivity whereas annotator 5 had 91.2% in sensitivity but at the cost of only 56.0% in specificity.

### Diagnostic Performance of APOLLAI Versus EMS

Comparison of sensitivity and specificity between EMS and APOLLAI against the ground truth was performed across the five neurological tasks (best gaze, facial palsy, motor arm, dysarthria, and aphasia) as well as for the global assessment.

APOLLAI demonstrated higher sensitivity than EMS across all neurological assessments. The greatest difference was observed for best gaze, for which sensitivity was markedly higher with APOLLAI than with EMS (93.3% versus 49.9%), with comparable specificity (89.7% versus 90.0%). Aphasia detection showed a similar pattern, with higher sensitivity for APOLLAI (71.9% versus 42.6%) and comparable specificity (88.4% versus 87.7%). For motor arm assessment, APOLLAI also demonstrated higher sensitivity than EMS (80.6% versus 58.7%), whereas EMS retained higher specificity (96.7% versus 90.6%). For dysarthria, APOLLAI likewise demonstrated higher sensitivity than EMS (80.5% versus 47.2%), with a modest reduction in specificity (90.1% versus 91.5%). Similarly, for facial palsy, APOLLAI demonstrated higher sensitivity than EMS (80.5% versus 51.3%), with a reduction in specificity (74.9% versus 89.2%). At the global level, APOLLAI demonstrated higher sensitivity than EMS (78.5% versus 59.4%) and slightly improved specificity (87.0% versus 85.9%).

For the global assessment, we performed a bootstrap analysis over 2000 iterations, following the methodology described in ^23^ to compute the confidence interval. The results are the following:

- SE(APOLLAI) – SE(EMS) = 20,6% [IC95%: +14.6%; +26.6%]
- SP(APOLLAI) – SP(EMS) = 0,2% [IC95%: −5.7%; +6.7%]

Bootstrap analysis over 2,000 iterations showed that APOLLAI significantly outperformed EMS in terms of sensitivity, with an absolute increase of 20.6% (95% CI: +14.6% to +26.6%). In contrast, the difference in specificity was minimal (+0.2%) and not statistically significant, as the corresponding 95% confidence interval included zero (−5.7% to +6.7%). Overall, these findings indicate that APOLLAI provides a marked improvement in sensitivity without compromising specificity.

### APOLLAI Discriminative Performance

APOLLAI demonstrated consistently high discriminative performance across all five neurological items, with AUC values ranging from 0.839 to 0.966 (Figure 2). Performance was highest for best gaze (AUC=0.966), followed by motor arm (AUC=0.913), dysarthria (AUC=0.911), and aphasia (AUC=0.877), whereas facial palsy showed the lowest discriminative performance (AUC=0.839). When combining all five neurological tasks, APOLLAI achieved a global AUC of 0.882, confirming preserved discriminative ability across the complete neurological examination.

**Figure 2.**
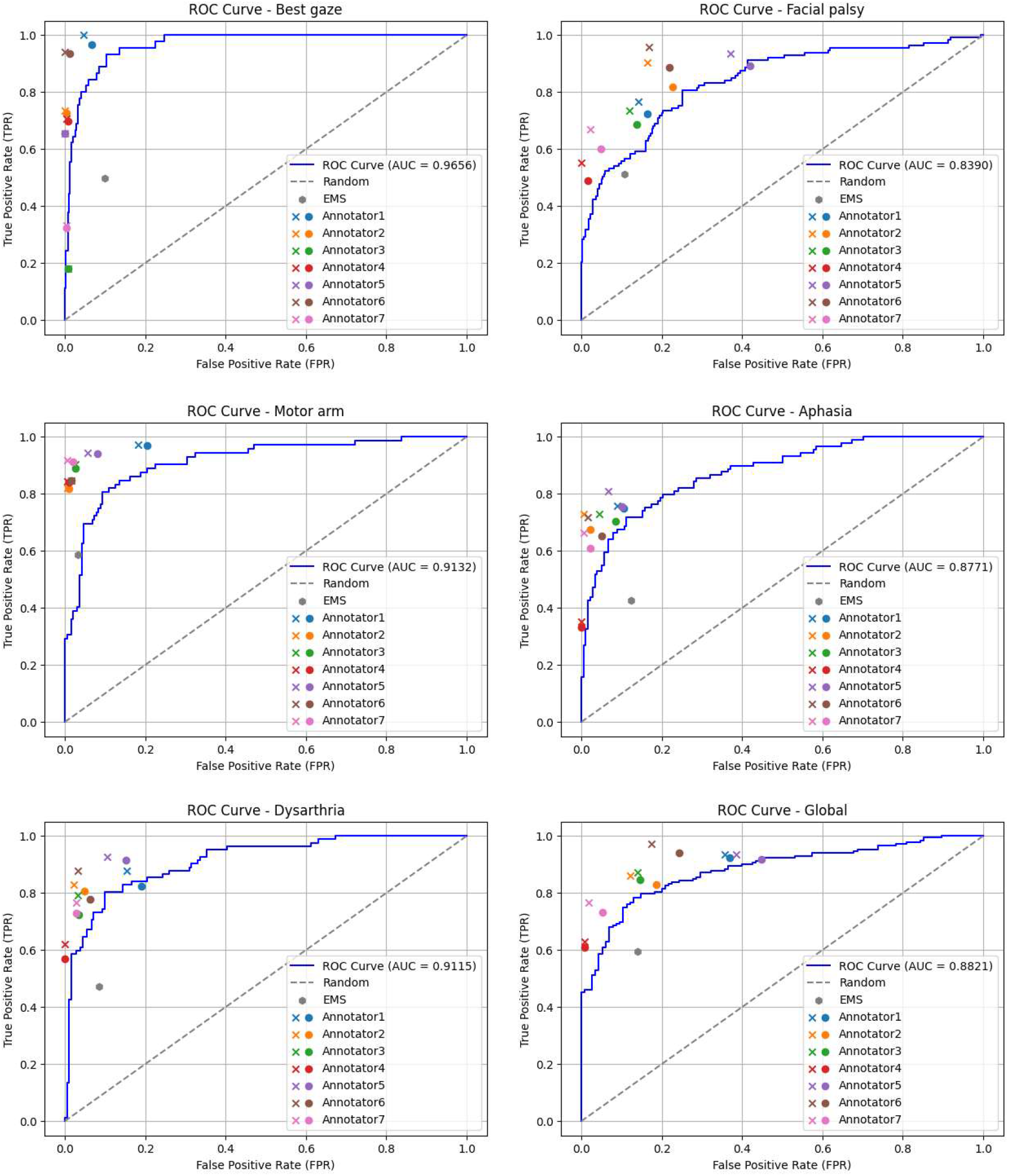
ROC curves of APOLLAI. Receiver operating characteristic curves showing the discriminative performance for each of the five neurological tasks as well as the combined global assessment. In all cases, ROC curves were generated using the consensus of the seven neurologists as the ground truth reference. Operating points corresponding to the consensus of the seven neurologists are represented by colored crosses (x), while individual neurologist operating points obtained using the OVR approach are shown as matching-colored circles (•). EMS operating points are displayed as gray hexagons. All operating points are shown relative to the APOLLAI ROC curves.

Individual neurologist operating points (using the OVR approach) were mostly positioned above the corresponding APOLLAI ROC curve, reflecting higher sensitivity than the model at comparable false-positive rates. However, a few points were positioned on or even below the curve, meaning that the AI model could perform as well as or even better than some individual neurologist in some cases.

The EMS operating point fell consistently below the APOLLAI ROC curve for four of the five items and for global assessment with the largest gap observed for best gaze and aphasia. The only exception was motor arm, where the EMS operating point lay close to the APOLLAI ROC curve itself.

## Discussion

### Inter-rater agreement for NIHSS tasks is only moderate

To our knowledge, this study is the first one to compute inter-rater agreement for 5 items of the NIHSS on a large scale with 435 subjects and seven NIHSS certified neurologists. The neurologists had no direct access to the patients but rather watched videos of them. Although this is a limitation of the study, the videos were specifically acquired for such a distant assessment. Having the seven annotators in the same room as the patient would have prevented them from having the same angle of vision; additionally, they would not have been fully blind to one another.

The observed interobserver agreement ranged from moderate to substantial. The pairwise Cohen’s κ values ranged from 0.17 to 0.81 (mean κ=0.523) and never reached excellent agreement, consistent with prior reports of substantial NIHSS interrater variability even among large, certified samples of clinicians^24^. This indicates that part of the diagnostic ceiling observed for both EMS and APOLLAI reflects an intrinsic difficulty of neurological assessment rather than a limitation specific to either approach.

In a previous paper about the same study^21^, the ground truth was the annotation of only one neurologist and not a consensus vote. For global assessment, we reported a sensitivity of 67.3% with a specificity of 81.5% for EMS. However, with the consensus vote of seven neurologists as new ground truth, the results are quite different: EMS sensitivity drops to 59.4%, and specificity is up to 85.9%. Assessing the NIHSS is a difficult task, even for a trained and NIHSS-certified neurologist: for research work, several annotators should assess the patients, and a consensus vote should serve as ground truth.

### APOLLAI consistently outperformed EMS staff

In this prospective study, APOLLAI demonstrated higher sensitivity than EMS personnel for the detection of five stroke-related neurological deficits (best gaze, facial palsy, motor arm, aphasia, and dysarthria) and for the global stroke assessment, while specificity remained broadly comparable between the two approaches for most tasks, with EMS retaining a notably higher specificity for motor arm and APOLLAI showing a slightly improved specificity at the global level (Table 3). Sensitivity and specificity values for APOLLAI were derived at an operating point on the ROC curve selected during model development; unlike human evaluators, who operate at a fixed sensitivity-specificity trade-off, APOLLAI offers the additional advantage of an adjustable operating point along the ROC curve, allowing the balance between sensitivity and specificity to be tailored to the clinical context. These findings directly contribute to recent calls for robust validation of AI-based decision-support tools in stroke care, which have so far been evaluated predominantly within hospital-based imaging workflows rather than under prehospital field conditions^25,26^.

**Table 3.** Sensitivity and specificity of EMS and APOLLAI for individual neurological tasks and global assessment. Sensitivity (SE) and specificity (SP) values for EMS and APOLLAI across the five neurological tasks (best gaze, aphasia, motor arm, dysarthria, and facial palsy) and for the combined global assessment.

|  | EMS |  | APOLLAI |  |
| --- | --- | --- | --- | --- |
|  | SE | SP | SE | SP |
| Best gaze | 49.9% | **90.0%** | **93.3%** | 89.7% |
| Facial palsy | 51.3% | **89.2%** | **80.5%** | 74.9% |
| Motor arm | 58.7% | **96.7%** | **80.6%** | 90.6% |
| Aphasia | 42.6% | 87.7% | **71.9%** | **88.8%** |
| Dysarthria | 47.2% | **91.5%** | **80.5%** | 90.1% |
| Global | 59.4% | 85.9% | **78.5%** | **87.0%** |

### Task level Analysis

The largest performance gap between APOLLAI and EMS was observed for best gaze (sensitivity, 93.3% versus 49.9%) and APOLLAI also achieved the highest discriminative performance for best gaze (AUC=0.966). Interestingly, there is growing clinical evidence that best gaze carries substantial diagnostic value beyond basic stroke recognition, including as a marker for large vessel occlusion in telemedicine-based assessment^27^, suggesting that this neurological sign, while underutilized in current prehospital practice, is highly amenable to objective, video-based detection.

For aphasia and dysarthria, a significant performance gap was also observed between APOLLAI and EMS, reflecting the nuanced linguistic judgment required to identify subtle language impairment under field conditions. Standard prehospital scales such as FAST and CPSS assess speech through a single undifferentiated item, without distinguishing aphasia, a language disorder, from dysarthria, a purely motor articulation deficit, which may compound the difficulty of detecting subtle impairment under field conditions.

Sensitivity for facial palsy was also higher for APOLLAI than for EMS (80.5% versus 51.3%), consistent with the pattern observed for the other four neurological tasks, though at the cost of the largest relative reduction in specificity among items (74.9% versus 89.2% for EMS). Despite this sensitivity gain, facial palsy showed the lowest discriminative performance for APOLLAI (AUC=0.839) among the five neurological tasks. This combination of genuine sensitivity gains at the selected operating threshold, alongside comparatively limited overall discrimination, is consistent with the inter-rater agreement findings reported above, where facial palsy showed the most consistent agreement across annotator pairs (lowest SD=0.076) but only a moderate mean κ (0.422). Together, these findings suggest that facial palsy assessment, whether performed by human raters or by APOLLAI, may be limited less by inconsistency between raters than by the intrinsically graded and subtle nature of the sign itself, which could reduce the separability of normal from abnormal presentations regardless of the assessment method.

Finally, APOLLAI displayed a higher sensitivity than EMS for the arm motor task (80.6% versus 58.7%) but with the second largest drop in specificity (90.6% versus 96.7%), and the point representing EMS’s performance for arm motor being slightly above the APOLLAI’s ROC curve, contrary to EMS points for all other tasks and for the global assessment. Specifically for the arm motor task, the video acquisition done during phase 1 led to difficulties analyzing the videos for APOLLAI: the positioning of the camera raised occlusion issues, the frequent backlighting making the videos even more challenging for APOLLAI to perform well on. In future work, it will be important to set a proper camera shooting angle and pay more attention to backlighting.

### Global assessment analysis

Beyond initial symptom recognition, EMS providers are central to an effective chain of stroke care, as accurate field assessment enables rapid transport and timely activation of hospital stroke response systems. The findings of this study suggest that APOLLAI could serve as a standardized adjunct to EMS clinical judgment in this setting, particularly for neurological domains that are under-recognized within current training frameworks, such as best gaze and language disturbance. Given that nearly one-third of strokes remain unidentified before hospital arrival^7–9^, a tool capable of flagging additional suspected neurological deficits without substantially compromising specificity could contribute to more consistent triage and prenotification to stroke centers.

Specificity remained broadly preserved for most tasks and was slightly improved at the global level, although motor arm and facial palsy showed a more pronounced reduction relative to EMS; this trade-off is particularly relevant from an implementation standpoint, as limiting false-positive stroke identification may support more confident EMS decisions regarding bypass transport to endovascular-capable comprehensive stroke centers, previously identified as a key lever for reducing time-to-treatment in severe stroke^14^.

### Strengths and limitations

This study has several strengths. First, EMS diagnostic performance was assessed across a large number of personnel, providing a strong and relevant human comparator.

Second, a probabilistic consensus method (Dawid–Skene algorithm^15^) was applied across seven independent, certified stroke neurologists to establish the reference standard. This approach provides a more robust ground truth than the single-rater annotations used in our earlier work^21^ and in most previous AI-based stroke detection studies. In addition, the OVR framework benchmarks each neurologist against the consensus of the remaining six raters rather than against an assumed infallible reference. Third, the ground truth used in this study was based on the clinical neurological examination rather than neuroimaging confirmation of stroke. This choice is consistent with the intended purpose of APOLLAI, which was designed and evaluated to approximate the bedside judgment of a neurologist. Consequently, APOLLAI should not be viewed as a substitute for CT or MRI, but rather as a tool that reproduces clinical neurological assessment.

Several limitations should be acknowledged. First, video recordings were acquired in a controlled hospital setting, typically several hours to days after stroke onset and following initiation of treatment, under conditions substantially different from those of acute prehospital encounters including reduced patient distress, standardized filming angles, and the absence of time pressure. This represents the most important limitation of the present study and must be considered when interpreting the reported performance estimates. Second, all patients included in this study were French-speaking, which may limit the generalizability of language-based assessments particularly for aphasia and dysarthria to populations speaking other languages. Third, this study was conducted within a single regional EMS system in southern France, and diagnostic performance may not generalize to systems with different training protocols, staffing models, or patient populations; multicenter validation is needed before broader conclusions can be drawn. Fourth, as in our earlier work^21^, EMS personnel were asked a global question regarding the presence of stroke signs and then asked to attribute observed abnormalities to specific tasks, rather than being asked targeted binary questions for each neurological domain; this may have introduced ambiguity, particularly between aphasia and dysarthria, which share overlapping speech-related video content.

## Conclusion

By incorporating a consensus-based reference standard derived from seven certified neurologists, this study further demonstrated that expert interobserver variability itself imposes meaningful limits on neurological assessment, reinforcing the relevance of evaluating diagnostic tools against multi-rater consensus rather than a single human reference. In this study, APOLLAI demonstrated higher sensitivity than EMS personnel for the detection of stroke-related neurological deficits, with specificity broadly preserved across tasks and at the global assessment level. These findings support the potential of APOLLAI as a standardized, scalable adjunct to prehospital stroke recognition, warranting prospective validation in real-world EMS practice. Future work should prioritize prospective, real-world field validation of APOLLAI across multiple EMS systems and geographic regions, using a dedicated mobile application integrated into routine prehospital workflows alongside standard scales such as FAST, CPSS, or BE-FAST. A prospective field study will start at the end of 2026 in the United States to acquire real-world prehospital data under live EMS conditions, which will enable assessment of APOLLAI performance across diverse operational contexts and represent a critical step toward demonstrating clinical utility beyond the controlled conditions of the present study.

## Data Availability

The datasets analyzed during the study are not publicly available because they contain sensitive patient data and are subject to privacy and ethical restrictions. The data cannot be shared outside the institution in accordance with applicable regulations and ethics approvals.

## Acknowledgments

The authors thank the Nîmes University Hospital (CHU de Nîmes) and its personnel for their participation in the data collection phase, and the Service Départemental d’Incendie et de Secours de l’Hérault (SDIS34) and its personnel for their participation in the stroke assessment sessions.

## Supplementary materials

As shown in Figure 3, the AUC varies considerably depending on which neurologist is excluded from the consensus. For aphasia, the AUC ranges from 0.8467 to 0.8814, while for dysarthria, it ranges from 0.8762 to 0.9218. In contrast, some tasks show very limited variability. For example, in the “Best Gaze” task, the difference between the lowest and highest AUC values is less than 0.01, indicating a much higher level of agreement among neurologists regarding the reference labels. These results are consistent with the inter-rater variability observed in the Kappa analysis and further underscore the absence of a single gold standard in neurological assessment. They demonstrate that performance estimates can be significantly influenced by the composition of the reference consensus, thereby highlighting the importance of pooling assessments from multiple experts to mitigate annotation uncertainty and establish a more reliable reference standard.

**Figure 3.**
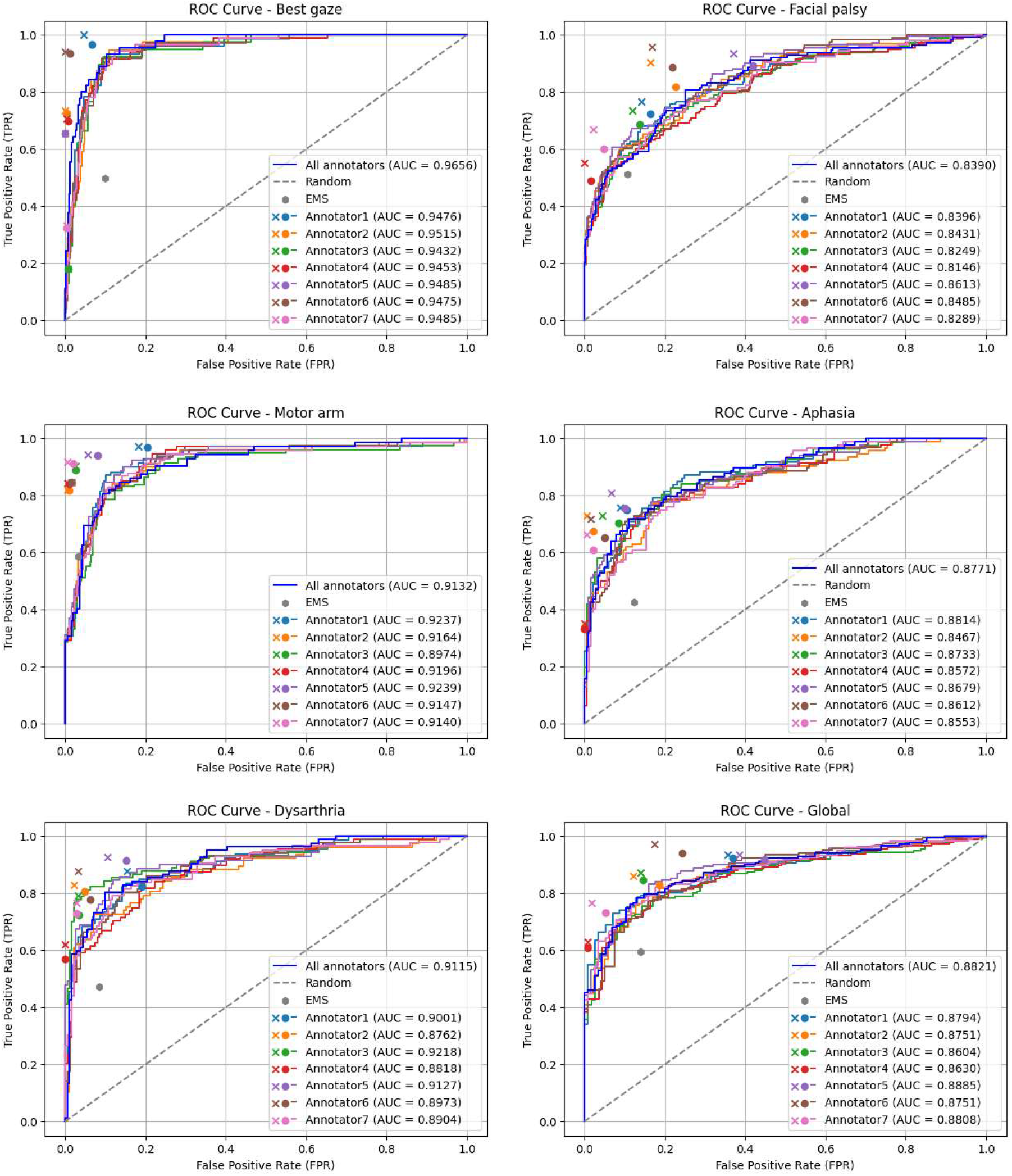
ROC curves of APOLLAI. Receiver operating characteristic (ROC) curves showing the discriminative performance of APOLLAI across the five neurological tasks and the combined global assessment. In each panel, the dark blue ROC curve was generated using the consensus of the seven neurologists as the reference standard. The other colored ROC curves correspond to APOLLAI models trained and evaluated using the different one-versus-rest (OVR) consensus. Colored crosses (x) denote the operating points of the seven-neurologist consensus, whereas colored circles (•) indicate the operating points derived from the OVR consensus. Gray hexagons represent EMS operating points. All operating points are shown in relation to the corresponding APOLLAI ROC curves.

## References

1. GBD 2016 Lifetime Risk of Stroke Collaborators, Feigin VL, Nguyen G, et al. Global, Regional, and Country-Specific Lifetime Risks of Stroke, 1990 and 2016. N Engl J Med. 2018;379(25):2429–2437. doi:10.1056/NEJMoa1804492

2. Katan M, Luft A. Global Burden of Stroke. Seminars in Neurology. Published online 2018. doi:10.1055/s-0038-1649503

3. GBD 2015 Neurological Disorders Collaborator Group. Global, regional, and national burden of neurological disorders during 1990-2015: a systematic analysis for the Global Burden of Disease Study 2015. Lancet Neurol. 2017;16(11):877–897. doi:10.1016/S1474-4422(17)30299-5

4. Krishnamurthi RV, Moran AE, Feigin VL, et al. Stroke Prevalence, Mortality and Disability-Adjusted Life Years in Adults Aged 20-64 Years in 1990-2013: Data from the Global Burden of Disease 2013 Study. Neuroepidemiology. 2015;45(3):190–202. doi:10.1159/000441098

5. Ovbiagele B, Goldstein LB, Higashida RT, et al. Forecasting the future of stroke in the United States: a policy statement from the American Heart Association and American Stroke Association. Stroke. 2013;44(8):2361–2375. doi:10.1161/STR.0b013e31829734f2

6. Saver JL. Time is brain--quantified. Stroke. 2006;37(1):263–266. doi:10.1161/01.STR.0000196957.55928.ab

7. Oostema JA, Chassee T, Baer W, Edberg A, Reeves MJ. Brief Educational Intervention Improves Emergency Medical Services Stroke Recognition | Stroke. Stroke. 2019;50(5):1193–1200. doi:10.1161/STROKEAHA.118.023885

8. Caffarelli M, Wood AJ, Crowe RP, et al. Prevalence and Determinants of Prehospital Impression of Stroke in Ischemic Stroke Patients. Neurology. Preprint posted online September 27, 2024. doi:10.1101/2024.09.25.24314407

9. Brandler ES, Sharma M, McCullough F, et al. Prehospital Stroke Identification: Factors Associated with Diagnostic Accuracy. J Stroke Cerebrovasc Dis. 2015;24(9):2161–2166. doi:10.1016/j.jstrokecerebrovasdis.2015.06.004

10. Harbison J, Hossain O, Jenkinson D, Davis J, Louw SJ, Ford GA. Diagnostic accuracy of stroke referrals from primary care, emergency room physicians, and ambulance staff using the face arm speech test. Stroke. 2003;34(1):71–76. doi:10.1161/01.str.0000044170.46643.5e

11. Kothari RU, Pancioli A, Liu T, Brott T, Broderick J. Cincinnati Prehospital Stroke Scale: reproducibility and validity. Ann Emerg Med. 1999;33(4):373–378. doi:10.1016/s0196-0644(99)70299-4

12. Llanes JN, Kidwell CS, Starkman S, Leary MC, Eckstein M, Saver JL. The Los Angeles Motor Scale (LAMS): a new measure to characterize stroke severity in the field. Prehosp Emerg Care. 2004;8(1):46–50. doi:10.1080/312703002806

13. Brott T, Adams HP, Olinger CP, et al. Measurements of acute cerebral infarction: a clinical examination scale. Stroke. 1989;20(7):864–870. doi:10.1161/01.STR.20.7.864

14. Chennareddy S, Kalagara R, Smith C, et al. Portable stroke detection devices: a systematic scoping review of prehospital applications. BMC Emerg Med. 2022;22(1):111. doi:10.1186/s12873-022-00663-z

15. Dawid AP, Skene AM. Maximum Likelihood Estimation of Observer Error-Rates Using the EM Algorithm. Journal of the Royal Statistical Society Series C (Applied Statistics). 1979;28(1):20–28. doi:10.2307/2346806

16. McHugh ML. Interrater reliability: the kappa statistic. Biochem Med (Zagreb). 2012;22(3):276–282. doi:10.11613/BM.2012.031

17. Nahm FS. Receiver operating characteristic curve: overview and practical use for clinicians. Korean J Anesthesiol. 2022;75(1):25–36. doi:10.4097/kja.21209

18. Hanley JA, McNeil BJ. The meaning and use of the area under a receiver operating characteristic (ROC) curve. Radiology. 1982;143(1):29–36. doi:10.1148/radiology.143.1.7063747

19. Landis JR, Koch GG. The measurement of observer agreement for categorical data. Biometrics. 1977;33(1):159–174. doi:10.2307/2529310

20. Cortes C, Vapnik V. Support-vector networks. Mach Learn. 1995;20(3):273–297. doi:10.1007/BF00994018

21. Pibre L, Yriarte F, Javault C. Enhancing pre-hospital stroke diagnosis with videos of the patient and AI: benchmarking AI against EMS personnel. Neuroscience Informatics. 2026;6(3):100277. doi:10.1016/j.neuri.2026.100277

22. Youden WJ. Index for rating diagnostic tests. Cancer. 1950;3(1):32–35. doi:10.1002/1097-0142(1950)3:1<32::aid-cncr2820030106>3.0.co;2-3

23. DiCiccio TJ, Efron B. Bootstrap Confidence Intervals. Statistical Science. 1996;11(3):189–212. doi:10.1214/ss/1032280214

24. Josephson SA, Hills NK, Johnston SC. NIH Stroke Scale reliability in ratings from a large sample of clinicians. Cerebrovasc Dis. 2006;22(5-6):389–395. doi:10.1159/000094857

25. Jalo H, Seth M, Pikkarainen M, et al. Early identification and characterisation of stroke to support prehospital decision-making using artificial intelligence: a scoping review protocol. BMJ Open. 2023;13(5):e069660. doi:10.1136/bmjopen-2022-069660

26. Elfahim O, Edjinedja KL, Cossus J, Youssfi M, Barakat O, Desmettre T. A Systematic Literature Review of Artificial Intelligence in Prehospital Emergency Care. Big Data and Cognitive Computing. 2025;9(9):219. doi:10.3390/bdcc9090219

27. Schröter N, Weiller A, Rijntjes M, et al. Identifying large vessel occlusion at first glance in telemedicine. J Neurol. 2023;270(9):4318–4325. doi:10.1007/s00415-023-11775-2

